# Association of Increased Fluvoxamine Use with Reports of Benefit for COVID

**DOI:** 10.1101/2022.01.19.22269020

**Authors:** N Kojima, Dima M Qato, JD Klausner

**Affiliations:** Department of Medicine, University of California Los Angeles, Los Angeles, 90095; Program on Medicines and Public Health, Department of Clinical Pharmacy, University of Southern California, School of Pharmacy, Los Angeles, California; Schaeffer Center for Health Policy and Economics, University of Southern California, Los Angeles, California; Department of Population and Public Health Sciences, University of Southern California, Keck School of Medicine, Los Angeles, 90033

**Keywords:** COVID-19, SARS-CoV-2, Vaccination, Prior infection

## Abstract

**Introduction:** Several recent studies have found that fluvoxamine decreases the risk of serious disease progression among people with early SARS-CoV-2 infection. In this study, we examined changes in the total number of fluvoxamine tablets dispensed across retail pharmacies in the U.S.

**Methods:** We hypothesized that fluvoxamine prescriptions would increase substantially since March 2021 as an option for the early treatment of SARS-CoV-2 infection. We used the IQVIA National Prescription Audit (NPA) Weekly nationally projected data for prescriptions dispensed from retail pharmacies for fluvoxamine from 27 December 2019 to 31 December 2021. We performed an interrupted-time series analyses on frequency of dispensing fluvoxamine tablets.

**Results:** The weekly rate of dispensed tablets of fluvoxamine increased throughout the study period. The weekly number of dispensed tablets of fluvoxamine increased (11.1%) from a baseline of 1,586,154 (95% confidence interval [CI]: 1,563,960 – 1,608,348) to 1,762,381 (95% CI: 1,735,682 – 1,789,080) by December 2021.

**Conclusion:** Our findings are consistent with a modest increase in the use of fluvoxamine for the treatment of COVID-19 associated with the discovery and media dissemination of the potential clinical benefit of fluvoxamine use.

## Main Text

Several recent studies have found that fluvoxamine decreases the risk of serious disease progression among people with early SARS-CoV-2 infection.^1-3^ In March 2021 a widely watched news program aired a report describing the potential benefit of fluvoxamine for COVID-19.^4^ While several medications were developed to treat COVID-19, repurposing existing medications like fluvoxamine offer benefits, including widespread availability, known safely profiles, and low cost, that newly approved medications lack.^5^ In this study, we examined changes in the total number of fluvoxamine tablets dispensed across retail pharmacies in the U.S. We hypothesized that fluvoxamine prescriptions would increase substantially since March 2021 as an option for the early treatment of SARS-CoV-2 infection.

We used the IQVIA National Prescription Audit (NPA) Weekly nationally projected data for prescriptions dispensed from retail pharmacies for fluvoxamine from 27 December 2019 to 31 December 2021. The NPA aggregates prescription data from pharmacies across the US representing approximately 92% of all retail prescription activity. We performed an interrupted-time series analyses on frequency of dispensing fluvoxamine tablets. The potential clinical benefit of fluvoxamine for COVID-19 was widely disseminated on 7 March 2021, therefore we compared periods from 27 December 2019 to 3 March 2021 and from 12 March to 31 December 2021. Adjustment for first-order autocorrelation was made though autoregressive integrated moving average models. The weekly number of dispensed tablets of fluvoxamine were compared between periods. Analyses were performed on StataSE (StataCorp, College Station, TX).

The weekly rate of dispensed tablets of fluvoxamine increased throughout the study period. The weekly number of dispensed tablets of fluvoxamine increased (11.1%) from a baseline of 1,586,154 (95% confidence interval [CI]: 1,563,960 – 1,608,348) to 1,762,381 (95% CI: 1,735,682 – 1,789,080) by December 2021 (Figure).

**Figure.**
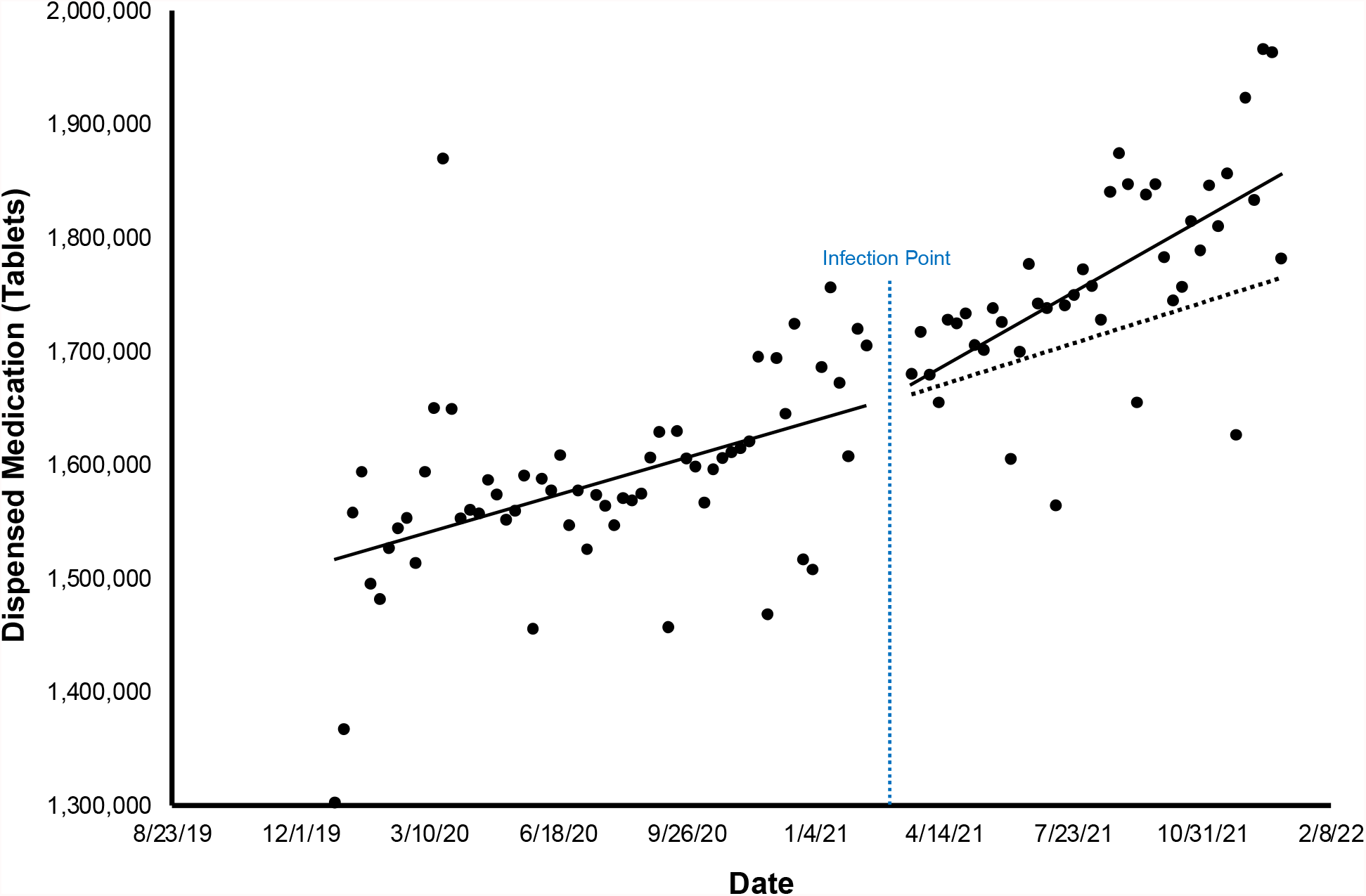
Trends in the total number of dispensed tablets of fluvoxamine in retail, mail, and long-term care pharmacies in the U.S., 27 December 2019 to 31 December 2021 *ARIMA regression of 3,234.2 (95% CI: 1,936.8 – 4,531.6; p<0.001).

Our findings are consistent with a modest increase in the use of fluvoxamine for the treatment of COVID-19 associated with the discovery and media dissemination of the potential clinical benefit of fluvoxamine use. Other events, however, may have contributed to this temporal trend. For instance, there may have also been increased fluvoxamine prescription for the treatment of depression, which is also associated with COVID-19. However, the increased use of fluvoxamine was non-linear and appeared to rapidly increase after increased awareness of its potential COVID-19 treatment benefit. Nevertheless, as an ecological study, our results are preliminary and should be confirmed with additional studies.

## Data Availability

All data produced in the present study are available upon reasonable request to the authors

## Declarations

Declaration of competing interests None

## Acknowledgements

This study was supported, in part, through the IQVIA Institute Human Data Science Research Collaborative. The statements, findings, conclusions, views, and opinions contained and expressed herein are not necessarily those of IQVIA or any of its affiliated or subsidiary entities

## References

1. Lenze EJ, Mattar C, Zorumski CF, et al. Fluvoxamine vs Placebo and Clinical Deterioration in Outpatients With Symptomatic COVID-19: A Randomized Clinical Trial. JAMA 2020;324(22):2292–2300. DOI: 10.1001/jama.2020.22760.

2. Seftel D, Boulware DR. Prospective Cohort of Fluvoxamine for Early Treatment of Coronavirus Disease 19. Open Forum Infect Dis 2021;8(2):ofab050. DOI: 10.1093/ofid/ofab050.

3. Reis G, Dos Santos Moreira-Silva EA, Silva DCM, et al. Effect of early treatment with fluvoxamine on risk of emergency care and hospitalisation among patients with COVID-19: the TOGETHER randomised, platform clinical trial. Lancet Glob Health 2022;10(1):e42–e51. DOI: 10.1016/S2214-109X(21)00448-4.

4. Alfonsi S. Finding a possible early treatment for COVID-19 in a 40-year-old antidepressant. 60 Minutes. CBS, 2021. (https://www.cbsnews.com/news/fluvoxamine-antidepressant-drug-covid-treatment-60-minutes-2021-03-07/).

5. Facente SN, Reiersen AM, Lenze EJ, Boulware DR, Klausner JD. Fluvoxamine for the Early Treatment of SARS-CoV-2 Infection: A Review of Current Evidence. Drugs 2021;81(18):2081–2089. DOI: 10.1007/s40265-021-01636-5.

